# Examining Health-Related Risk Factors for Stroke and the Role of Health Insurance in Moderating Lifestyle: Evidence from the BRFSS

**DOI:** 10.1101/2025.10.08.25337014

**Authors:** Jinbo Niu

## Abstract

Stroke remains one of the leading causes of mortality and disability in the United States. However, there are still disparities in how lifestyle and healthcare access shape stroke outcomes. This study investigates the associations between smoking, alcohol consumption, body mass index (BMI), mental and physical health, sleep duration, health insurance status, and stroke incidence using data from the 2022 Behavioral Risk Factor Surveillance System (BRFSS, N = 77,146). Logistic regression models are applied to estimate the direct effects of these factors on stroke risk, as well as the moderating role of health insurance.

The results indicate that while some traditional risk factors, such as smoking and alcohol use, show weaker or unexpected associations with stroke in this dataset, mental health, physical health, and health insurance coverage demonstrate significant protective effects. In particular, interaction analyzes reveal that health insurance strengthens the beneficial impact of better mental and physical health and adequate sleep on reducing stroke risk. These findings highlight the importance of considering access to healthcare not only as an independent determinant of health, but also as a moderator that amplifies the benefits of healthy lifestyles.

This study contributes to public health research by underscoring the complex interaction between behavioral risk factors and structural determinants such as health insurance. Policies that expand access to affordable healthcare should improve the effectiveness of lifestyle-based prevention strategies and help reduce stroke incidence among disadvantaged populations.

## Introduction

Stroke has been identified as a significant public health concern and it is one of the leading causes of mortality and disability globally[1]. According to most studies, stroke is still the primary cause of mortality in the US and has been placing burdens on healthcare systems [2]. Therefore, it is crucial to identify potential risk factors and investigate their association with stroke. In previous studies, smoking, alcohol consumption, mental and physical health problems, and unhealthy Body Mass Index (BMI) have already been identified as the main attributes of stroke[3]. However, neither the extent of these variables in stroke nor their interaction effect has been pervasively studied in the population-level research literature.

The purpose of this study is to investigate the associations between the health-related factors mentioned above and stroke occurrence using a logistic regression model. The study aims to determine the extent to which these characteristics are associated. Furthermore, the study intends to investigate the possible interaction effects of health insurance status and how health insurance interacts with other variables. Understanding this interaction is critical to figuring out how the availability of healthcare interacts with other stroke risk factors. This research is based on data from the Centers for Disease Control and Prevention (CDC) Behavioral Risk Factor Surveillance System (BRFSS), an annual health survey that collects detailed health information from over 400,000 adults in the United States. The BRFSS collects data on health behaviors, chronic illnesses, and socioeconomic factors. The rich data types included in the dataset make it a valuable resource for studying public health concerns. Although most previous studies have employed the BRFSS to investigate various health outcomes, this study uniquely focuses on the interaction of health insurance status and lifestyle factors. It intends to investigate how healthcare access affects stroke preventive efforts.

However, disparities still exist in stroke outcomes across different populations even though there has been progress in stroke prevention [4]. The disparity is particularly prevalent among those with limited access to healthcare [5]. Also, socioeconomic factors such as access to health insurance have a significant impact on health outcomes[6]. People without health insurance may encounter difficulties getting preventative care or early stroke therapies [7]. Understanding how health insurance modifies the association between risk factors and stroke can aid in the development of a more effective intervention plan to minimize stroke incidence in disadvantaged populations.

By employing the logistic regression model, this study aims to answer the following research questions:

- What potential relationships exist among smoking, alcohol consumption, mental and physical health, sleeping hours, BMI, health insurance, sex and stroke?
- Is there an interaction effect between health insurance status and other variables?

These findings will help to deepen our understanding of the relationship between stroke risk variables and emphasize the relevance of health equality in stroke prevention initiatives. Identifying the interaction effects of health insurance may lead to more effective public health measures for lowering stroke rates in marginalized populations.

## Method

The purpose of this study is to look at the potential relationships between health-related characteristics (cigarette usage, alcohol intake, BMI, mental and physical health, sleeping hours, health insurance, and sex) and stroke incidence. Meanwhile, the study seeks to determine whether health insurance status can interact with the rest of the variables. To achieve these objectives, secondary data analysis will be carried out using the BRFSS data.

## Data

The data used in this study were obtained in 2022 by landline and mobile phone surveys in all 50 states, the District of Columbia, Guam, Puerto Rico, and the United States Virgin Islands [8]. The landline respondents were selected through the disproportionate stratified sampling (DSS) method, while cell phone respondents were chosen randomly, and each person had an equal chance of being chosen. The dataset has 328 variables and a total of 445,132 observations. Missing values are denoted by “NA.” The data were retrieved from the CDC’s official website and saved in SAS format.

Because the data was obtained using landline and cell phone surveys, various biases such as non-response, decline to answer, and incomplete interviews are possible according to [9]. To reduce potential bias, responses such as *blank, decline to answer*, or *Not sure/Don’t know* will be removed.

## Variables

From the 328 variables included in the dataset, eight are considered in the analysis:

- *Smoke*: Smoke at least 100 cigarettes (0 = No, 1 = Yes)
- *Alcohol*: Average alcoholic beverages per day over the past 30 days
- *BMI*: body mass index (BMI)
- *Mental*: Number of Days Mental Health Not Good per month (0-31)
- *Physical*: Number of Days Physical Health Not Good per month (0-31)
- *Sleeping*: How many hours to sleep per day (0-24)
- *Insurance*: Have any health insurance or not (0 = No, 1 = Yes)
- *Sex*: Male or Female
- *Stroke*: Ever diagnosed with stroke or not (0 = No, 1 = Yes)

### Analysis

All data analyses will be conducted using R software (R Foundation for Statistical Computing). Descriptive statistics will first be performed to summarize all the variables. To avoid multicollinearity, an initial exploratory data analysis (EDA) will be performed to assess the correlation between the dependent and independent variables, as well as the correlation between the independent variables.

Following the fundamental EDA, a logistic regression model will be constructed to investigate several independent variables. Odds ratios (OR), confidence intervals (CI), and p-values will be applied to evaluate the extent of association and the statistical significance of each variable.

To further explore the moderating role of health insurance on the relationships between the rest of the variables and the risk of stroke, interaction effects were included in the logistic regression models. Interaction terms were established between health insurance and each of the independent variables: *Smoke, Alcohol, Mental, Physical,Sleeping, BMI*, and *Sex*.

These interactions were explored to see if the effect of each factor on stroke risk is influenced by health insurance status. The significance of these interaction effects was determined by analysis of variance (ANOVA). A significant interaction implies that the relationship between the independent variable and stroke can be different if the health insurance status changes.

## Results

### Descriptive Statistics

The dataset used for analysis includes 77,146 completed records, with 2.92% reporting a history of stroke. The sample had a smoking prevalence of 40.98% and an average daily alcohol consumption of 2.4 drinks. Respondents had an average of 10.02 days of poor mental health and 4.69 days of poor physical health in the previous 30 days. The average sleep duration was 6.84 hours per day.

In terms of health coverage, 94.69% had health insurance. The sample was fairly balanced by gender, with 44.48% male and 55.52% female participation. These numbers offer a diverse sample of health behaviors and outcomes and enable further investigation of stroke risk and associated factors.

### Bivariate Analysis

The biserial correlations between the independent variable and the dependent variable were calculated to assess the degree of relationship between the two variables. The variable with the strongest connection is *Physical* (r=0.131), suggesting that poor physical health may be slightly connected to stroke. The biserial correlation results show weak connections between the continuous variables and *Stroke*. The correlations between *Mental, Alcohol, Sleeping*, and *BMI* are weak or near zero. Given the weak correlations, the continuous factors are unlikely to be strong individual predictors of *Stroke*. This emphasizes the need for a multivariate logistic model, as well as consideration of the potential interaction effect.

Table 2 presents the relationships between the five continuous variables: *Alcohol, Mental, Physical, Sleeping*, and *BMI*. The Spearman correlation coefficient has been applied and the coefficients range from −0.15 to 0.23, with the strongest being between *Mental* and *Physical* (0.23). This shows that the variables are generally independent of one another. As a result, multicollinearity is not an issue in the following multivariate logistic regression.

**Table 1:** Biseral Point Correlations for Various Variables.

| Variable | Biserial Point |
| --- | --- |
| Alcohol | 0.008 |
| Mental | 0.058 |
| Physical | 0.13 |
| Sleeping | -0.009 |
| BMI | 0.018 |

**Table 2:** Correlation matrix of continuous variables.

| Variable | Alcohol | Mental | Physical | Sleeping | BMI |
| --- | --- | --- | --- | --- | --- |
| Alcohol | 1.0000 | 0.1042 | -0.0100 | -0.0560 | 0.0293 |
| Mental | 0.1042 | 1.0000 | 0.2295 | -0.1548 | 0.0575 |
| Physical | -0.0100 | 0.2295 | 1.0000 | -0.0640 | 0.0865 |
| Sleeping | -0.0560 | -0.1548 | -0.0640 | 1.0000 | -0.0774 |
| BMI | 0.0293 | 0.0575 | 0.0865 | -0.0774 | 1.0000 |

**Table 3:** Chi-squared Test Results.

| Variable | $\chi^2$ | p |
| --- | --- | --- |
| Cigarette Use | 383.276 | <0.001 |
| Health Insurance | 4.478 | 0.034 |
| Sex | 10.848 | <0.001 |

Associations between nominal variables (*Smoke, Health, Insurance*) and the outcome variable (*Stroke*) have been identified based on the chi-squared tests. All these variables have been found associated with the outcome (p*<*0.001, p = 0.034, p *<*0.001, respectively).

### Multivariate Logistic Regression

In this section, logistic regression was used to look at the relationship between lifestyle factors and stroke incidence. The study indicated that those who have smoked more than 100 cigarettes have a significantly decreased risk of stroke (OR = 0.524, *p <*0.001). This contradicts previous research which has consistently shown that smoking increases stroke risk[10]. Alcohol use was associated with a slightly higher risk of stroke (OR = 1.01), but the effect was not statistically significant (*p* = 0.185). The p-value indicates that *Alcohol* has no substantial impact on stroke risk in this analysis. This finding also contradicts previous research which suggests less alcohol use came with lower stroke risk [11]. Both mental and physical health had small but statistically significant protective effects against stroke, with fewer unhealthy days associated with a lower likelihood of stroke (*Mental* OR = 0.991, *p <*0.001; *Physical* OR = 0.950, *p <*0.001). Despite research indicating the good effects of sleep on health [12], sleep duration did not reveal a significant influence on stroke risk (OR = 0.991, *p* = 0.499). BMI was not statistically significant (OR = 1.000, *p* = 0.211), indicating that body mass index does not influence stroke risk in this sample. Insured persons had a significantly lower stroke risk (OR = 0.696, *p* = 0.0007), highlighting the relevance of access to healthcare resources in reducing stroke incidence. As for sex, Males have about 11% lower odds of the outcome compared to females, and this difference is statistically significant (OR = 0.8913, *p* = 0.0091).

### Interaction effect

In this part, it will be determined whether the effect of having health insurance can be mitigated by other factors. New models with interactions will be created and compared to the first logistic model using ANOVA. Table 4 compares the results of ANOVA between the initial logistic model and the model with interactions. According to the ANOVA results of interaction effects, certain variables have statistically significant interactions with health insurance status. Both the interactions of *Mental* and the interaction of *Physical* with health insurance are statistically significant (p = 0.023 and p = 0.034, respectively). This suggests that these variables can have a different impact on stroke risk when they are combined with health insurance. Furthermore, the interaction effect of *Sleeping* is extremely significant (p = 0.001). This indicates that sleeping hours and health insurance status play a vital role in determining stroke risk. However, interactions between *Smoke, Alcohol, BMI* (body mass index), and *Sex* (gender) did not have statistical significance, indicating that their combined effects with health insurance are less influential in this study’s context.

**Table 4:** Logistic Regression Results.

| Variable | OR | CI Lower | CI Upper | p-value |
| --- | --- | --- | --- | --- |
| Smoke | 0.5236 | 0.4789 | 0.5721 | <0.001** |
| Alcohol | 1.0116 | 0.9954 | 1.0300 | 0.1852 |
| Mental | 0.9910 | 0.9867 | 0.9953 | <0.001** |
| Physical | 0.9497 | 0.9461 | 0.9534 | <0.001** |
| Sleeping | 0.9912 | 0.9661 | 1.0172 | 0.4990 |
| BMI | 1.0000 | 0.9999 | 1.0000 | 0.2106 |
| Insurance | 0.6958 | 0.5608 | 0.8531 | 0.0007** |
| Sex | 0.8913 | 0.8174 | 0.9719 | 0.0091** |

1. Significant $p$ -values ( $p < 0.05$ ) are marked by \*\*. 2. The reference group of smoke is 0 (No). 3. The reference group of Insurance is 0 (No). 4. The reference group of Sex is 0 (Female). 5. OR - Odds Ratio, CI = Confidence Interval.

**Table 5:** Anova outcomes between the initial model and models with interactions.

| **Interaction** | **Deviance** | **p** |
| --- | --- | --- |
| SMOKE100 | 2.0113 | 0.156 |
| AVEDRNK3 | 1.3946 | 0.238 |
| MENTHLTH | 5.1657 | 0.023** |
| PHYSHLTH | 4.5032 | 0.034** |
| SLEPTIM1 | 10.173 | 0.001** |
| BMI | 0.4111 | 0.521 |
| SEX | 0.3631 | 0.547 |

*Note: Significant p-values ( $p < 0.05$ ) are marked by \*\*.*

## Discussion

This study aimed to examine the relationships between lifestyle factors and stroke risk, with a special emphasis on whether health insurance status modifies these associations. The logistic model used on the BRFSS dataset allowed for the assessment of both direct effects and interactions related to stroke risk, providing insights into how these lifestyle factors influence stroke outcomes.

The findings demonstrated that several variables can significantly influence stroke risk. Contrary to most literature that identifies smoking as a major risk factor for stroke [10], smoking in this sample was unexpectedly found to be negatively associated with stroke. This anomaly may be a result of self-reported bias or limitations in the smoking measurement (i.e., ever smoked at least 100 cigarettes). Such bias can potentially result in an inaccurate capture of current smoking behaviors. Similarly, No significant association was found between alcohol consumption and stroke risk in this analysis and this is also contrary to literature that identifies alcohol consumption as a risk factor[13].

Both mental and physical health, as measured by the number of unwell days in the previous month, demonstrated minor but statistically significant protective effects against stroke. This implies that persons who have fewer days of poor mental or physical health are at a lower risk of stroke, underlining the importance of general well-being in stroke prevention. Sleeping hours also showed a protective impact, with longer sleep hours reducing stroke risk. The findings are consistent with existing research[14] demonstrating that proper sleep benefits cardiovascular health.

Notably, health insurance had a robust protective effect against stroke risk. Such an effect highlighted the critical role of healthcare access. People having insurance were considerably less likely to have a stroke. This could be attributed to improved access to preventive services such as regular health screenings and early detection of cardiovascular disease [15]. The interaction effect evaluation also revealed that health insurance can moderate the effects of mental health, physical health, and sleeping hours on stroke risk. Specifically, insured individuals with fewer poor mental or physical health days or longer sleep duration had an even lower stroke risk. This implies that access to healthcare magnifies the benefits of good health and enough rest.

However, the interaction effects of health insurance on smoking, alcohol consumption, BMI, and sex were not statistically significant. This suggests that healthcare access may have a more indirect influence on these behaviors and the influence is likely to be caused by broader social or personal factors beyond insurance coverage.

This study’s strengths include its large, representative sample and the exploration of interaction effects with a focus on the moderating role of health insurance. However, some limitations must be acknowledged. The reliance on self-reported data introduces potential biases, especially regarding sensitive behaviors like smoking and alcohol use. Additionally, the cross-sectional design of the BRFSS limits causal inferences about the relationships between lifestyle factors and stroke risk. Future longitudinal research could delve deep into these associations over time. Additionally, the logistic regression model can be applied to interpret the relationship between the dependent variable and the independent variable, but it is not capable of predicting the occurrence of stroke accurately based on the independent variables above due to the imbalanced data. Alternative prediction algorithms, such as random forest, as well as strategies for dealing with imbalanced data, such as the Synthetic Minority Over-sampling Technique (SMOTE), can be used to improve prediction[16].

## Conclusion

This study provides valuable insights into the relationship between lifestyle factors and stroke risk, with a focus on how health insurance status moderates these associations. The findings indicate that smoking, alcohol consumption, mental and physical health, sleep duration, and health insurance significantly influence stroke outcomes. Health insurance can enhance the protective effects of good mental health, physical health, and adequate sleep. This interaction effect highlights the crucial role of healthcare access in reducing stroke risk. While interactions with smoking, alcohol consumption, BMI, and sex were not significant. The results emphasize the need for further research to explore these complex relationships. Overall, this study highlights the importance of addressing both lifestyle behaviors and healthcare access in stroke prevention strategies. To minimize the risk of stroke and improve cardiovascular health outcomes, future initiatives should focus on promoting healthy behaviors and increasing access to healthcare for disadvantaged people.

## Data Availability

All data produced are available online at: https://www.cdc.gov/brfss/index.html

https://www.cdc.gov/brfss/index.html

## References

[1] M. Katan and A. Luft. Global burden of stroke. Seminars in Neurology, 38(2):208–211, 2018.

[2] A. K. Boehme, C. Esenwa, and M. S. Elkind. Stroke risk factors, genetics, and prevention. Circulation Research, 120(3):472–495, 2017.

[3] Y. Kono, Y. Terasawa, K. Sakai, Y. Iguchi, Y. Nishiyama, C. Nito, S. Suda, K. Kimura, Y. Murakami, T. Kanzawa, K. Yamashiro, R. Tanaka, and S. Okubo. Association between living conditions and the risk factors, etiology, and outcome of ischemic stroke in young adults. Internal medicine (Tokyo, Japan), 62(19):2813–2820, 2023.

[4] R. L. Sacco. Stroke disparities: From observations to actions: Inaugural edward j. kenton lecture 2020. Stroke, 51(11):3392–3405, 2020.

[5] S. C. O. Martins, T. L. Secchi, C. Molina, and R. Nogueira. Editorial: Development of stroke systems of care across the globe. Frontiers in Neurology, 14:1292036, 2023.

[6] D. J. McMaughan, O. Oloruntoba, and M. L. Smith. Socioeconomic status and access to healthcare: Interrelated drivers for healthy aging. Frontiers in Public Health, 8:231, 2020.

[7] M. R. MacDonald, S. Zarriello, J. Swanson, N. Ayoubi, R. Mhaskar, and A. S. Mirza. Secondary prevention among uninsured stroke patients: A free clinic study. SAGE Open Medicine, 8:2050312120965325, 2020.

[8] Centers for Disease Control and Prevention. Brfss, 2023. Retrieved Dec 20, 2023, from https://www.cdc.gov/brfss/index.html.

[9] G. Pariyo, A. Meghani, D. Gibson, J. Ali, A. Labrique, I. A. Khan, G. M. A. Kibria, H. Masanja, A. A. Hyder, and S. Ahmed. Effect of the data collection method on mobile phone survey participation in bangladesh and tanzania: Secondary analyses of a randomized crossover trial. JMIR Formative Research, 7:e38774, 2023.

[10] B. Pan, X. Jin, L. Jun, S. Qiu, Q. Zheng, and M. Pan. The relationship between smoking and stroke: A meta-analysis. Medicine, 98(12):e14872, 2019.

[11] A. I. Christensen, B. G. Nordestgaard, and J. S. Tolstrup. Alcohol intake and risk of ischemic and haemorrhagic stroke: Results from a mendelian randomisation study. Journal of Stroke, 20(2):218–227, 2018.

[12] U. Albakri, E. Drotos, and R. Meertens. Sleep health promotion interventions and their effectiveness: An umbrella review. International journal of environmental research and public health, 18(11):5533, 2021.

[13] Andrew Smyth, Martin O’Donnell, Sumathy Rangarajan, Graeme J. Hankey, Shahram Oveisgharan, Michelle Canavan, Clodagh McDermott, Denis Xavier, Hongye Zhang, Albertino Damasceno, Alvaro Avezum, Nana Pogosova, Aytekin Oguz, Danuta Ryglewicz, Helle Klingenberg Iversen, Fernando Lanas, Annika Rosengren, Salim Yusuf, Peter Langhorne, and on behalf of the INTERSTROKE Investigators. Alcohol intake as a risk factor for acute stroke. Neurology, 100(2):e142–e153, 2023.

[14] B. H. Huang, B. del Pozo Cruz, A. Teixeira-Pinto, et al. Influence of poor sleep on cardiovascular disease-free life expectancy: A multi-resource-based population cohort study. BMC Medicine, 21(1):75, 2023.

[15] L. N. Medford-Davis, G. C. Fonarow, D. L. Bhatt, H. Xu, E. E. Smith, R. Suter, E. D. Peterson, Y. Xian, R. A. Matsouaka, and L. H. Schwamm. Impact of insurance status on outcomes and use of rehabilitation services in acute ischemic stroke: Findings from get with the guidelines-stroke. Journal of the American Heart Association, 5(11):e004282, 2016.

[16] Y. Wu and Y. Fang. Stroke prediction with machine learning methods among older chinese. International Journal of Environmental Research and Public Health, 17(6):1828, 2020.

